# Language Matters -Representations of ‘heart failure’ in English discourse: a large-scale linguistic study

**DOI:** 10.1101/2022.02.10.22270750

**Authors:** Jane Demmen, Nick Hartshorne-Evans, Elena Semino, Rajiv Sankaranarayanan

## Abstract

**Aims:** Heart failure (HF) has a lower public profile compared to other serious health conditions, notably cancer. This discourse analysis study investigates the extent to which HF is discussed in general contemporary English, UK parliamentary debates, and the ways in which HF is framed in discussions, when compared to two other serious health conditions, cancer and dementia.

**Methods:** The Oxford English Corpus of 21^st^ century English-language texts (2 billion words) and the UK Hansard Reports of parliamentary debates from 1945 to early 2021 were used to investigate the relative frequencies, contexts of use of the terms ‘heart failure’, ‘cancer’ and ‘dementia’.

**Results:** In the Oxford English Corpus, the term ‘heart failure’ occurs 4.26 times per million words (pmw), ‘dementia’ occurs 3.68 times pmw and ‘cancer’ occurs 81.96 times pmw. Cancer is talked about 19 times more often than HF and 22 times more often than dementia. These are disproportionately high in relation to actual incidence: annual cancer incidence is 1.8 times that of the other conditions; annual cancer mortality is twice that caused by coronary heart disease (including heart failure) or dementia.

‘Heart failure’ is used much less than ‘cancer’ in UK parliamentary debates(House of Commons and House of Lords) between 1945 and early 2021, and less than ‘dementia’ from 1990 onwards. Moreover, HF is even mentioned much less than potholes in UK roads and pavements. In 2018, for example, ‘pothole/s’ were mentioned over 10 times pmw, 37 times more often than ‘heart failure’, mentioned 0.28 times pmw. Discussions of HF are comparatively technical and formulaic, lacking the survivor narratives that occur in discussions of cancer.

**Conclusions:** HF is under-discussed in contemporary English compared to cancer and dementia. HF is also under-discussed in UK parliamentary debates, even compared to the less-obviously life threatening topic of potholes in roads and pavements.

**What is already known on this topic:** Heart failure is a serious health condition with significant morbidity and mortality, which is comparable to other serious health conditions such as cancer.

**What this study adds:** Our study has shown that heart failure is less frequently discussed in contemporary English as well as in UK parliamentary debates in comparison to other serious health conditions such as cancer and dementia, despite comparably significant adverse outcomes and also that discussions regarding people with heart failure are less empowering in comparison to discussions regarding cancer.

**How this study might affect research, practice or policy?:** Results of this study should motivate all stakeholders involved in heart failure to redouble their efforts to spread awareness regarding the seriousness of the condition in general discourse as well as to engage parliamentarians better and thereby exert influence upon commissioners to significantly improve investment in prevention, early diagnosis and better management of heart failure.

## Introduction

Heart failure (HF) is a significant public health issue with an estimated global prevalence of sixty five million^1^, although the prevalence of known HF in the western world is around 1-2%^2^. The prevalence of HF is predicted to increase significantly due to the ageing population, as well as improved survival from other medical conditions such as ischaemic heart disease, hypertension and disease^1^. HF leads to high morbidity through poor health related quality of life^3^ and recurrent hospitalisations with a 30 day readmission rate of around 20%^4^. Heart failure also contributes to increased mortality (30 day, 1, 2, 5 and 10-year survival to be 10%, 20%, 27%, 43% and 65%, respectively^5-7^. Heart failure consumes 1-2% of the annual healthcare budget in Europe and USA ^8,9^ with the majority of costs (>70% directed towards hospital care). Studies have shown that mortality due to HF is worse than certain types of cancer^10,11^. However, HF has not received a similar priority or profile such as other serious health conditions like as cancer in terms of government policy or funding and thus cancer has seen a much greater improvement in survival^12,13^. The improvement in cancer survival rates has been attributed to improvements in diagnosis and treatment due to better investment as well as changes in infrastructure since the introduction of the Cancer Plan in the UK two decades ago^14^

Large-scale discourse analysis using computer-assisted methods has been shown to be useful to understanding how people think and feel about serious health conditions, including cardiovascular disease and cancer^15,16^. There is little analysis to date of the way HF, specifically, is represented, apart from Strong and Gilmour’ s study of internet texts^17^. They noted that biomedical discourses (of a medical/technical nature, such as we found) were dominant, but also noted narratives of ‘living with heart failure’ which were scarce in our data. They noted an absence of talk about the contribution of nurses and the “emotional and spiritual dimensions of heart failure”, which were also not noted in our data. We therefore conducted this study to investigate the extent to which HF is discussed in general contemporary English as well UK parliamentary debates, and in particular, compare reference to HF with discussions about other serious health conditions such as cancer and dementia. We also compare the frequency of references to HF in UK Parliamentary debates with references to a non-medical topic, namely, pot-holes on UK roads.

## Methods

The study was commissioned by the Pumping Marvellous Foundation, a UK HF patient charity funded by donations and fundraising by individuals, with support from the NHS and charitable organisations plus corporate sponsorship. The study was conducted by linguists at the ESRC Centre for Corpus Approaches to Social Science, a research centre at Lancaster University which specialises in applying computer-assisted frequency-based statistical methods to the study of language in social life using large bodies of text.

Use of the terms ‘heart failure’ and ‘cancer’ was investigated in the following two data sets, or ‘corpora’ :

- The Oxford English Corpus (OEC): 2,073,319,589 words of contemporary (21^st^ century) English from the UK, US, Ireland, Australia, New Zealand, the Caribbean, Canada, India, Singapore, and South Africa, compiled by Oxford Languages (Oxford University Press). Its contents are sourced mainly from web-based material supplemented by some printed texts and are grouped into the genres Medicine, News, Fiction, Life and leisure, Science, Society, Weblog, Arts, Sport, Business, Religion, Humanities, Law, Military, Computing, Agriculture, Environment, Paranormal, Transport, Games plus an Unclassified category. The OEC is accessible by subscription through SketchEngine^18^ (Lexical Computing), a web-based interface providing access to a range of corpora and corpus linguistics software tools (https://www.sketchengine.eu/).
- The Hansard Corpus (HC): Hansard reports of parliamentary debates in UK House of Commons & Lords from 1 January 1945 up to and including 25 February 2021, accessed through Hansard at Huddersfield, a publicly-accessible interface and search tool provided by the University of Huddersfield (https://hansard.hud.ac.uk/site/site.php)^19^. More recent debates were accessed through the UK Parliament Hansard website (hansard.parliament.uk).

We refer to ‘heart failure’, ‘cancer’ and ‘dementia’ as linguistic terms when cited in single quotation marks, and as illnesses when not in quotation marks.

### Statistical analysis

Simple frequency counts of occurrences of linguistic terms were carried out automatically by the software in the SketchEngine interface. SketchEngine also identified words which tend to co-occur most typically (‘collocates’) by computing LogDice^20^scores measuring the strength of relationships between words and displaying these in rank order from most to least typical.

## Results

### Comparative frequency of use of the terms ‘heart failure’, ‘cancer’ and ‘dementia’ in contemporary English

Table 1 shows the actual (raw) frequencies of use for each term in the whole data set of contemporary English in the OEC (*n*) and the relative frequency per million words (pmw).

**Table 1.** Comparison of relative frequency of ‘heart failure’, ‘cancer’ and ‘dementia’ in contemporary English

| **‘heart failure’** |  | **‘cancer’** |  | **‘dementia’** |  |
| --- | --- | --- | --- | --- | --- |
| *n* | pmw | *n* | Pmw | *n* | pmw |
| 10,350 | 4.26 | 199,251 | 81.96 | 8,945 | 3.68 |
| pmw = per million words |  |  |  |  |  |

Across all geographical varieties of contemporary English the term ‘heart failure’ was mentioned much less often than the term ‘cancer’. The greatest disparity was in Irish English, where ‘cancer’ was mentioned 111 times more often than ‘heart failure’, and the least disparity was in American English where ‘cancer’ was mentioned 14 times more often. In British English ‘cancer’ was mentioned 22 times more often than ‘heart failure’. The comparison with ‘dementia’ was a little less consistent. ‘Heart failure’ was mentioned less often than ‘dementia’ in all varieties of English except for American English, where it was mentioned about twice as often, and East Asian English (about one and a half times more often). In British English the terms were mentioned with quite similar frequency (‘heart failure’ nearly 4.5 times pmw and ‘dementia’ just over 5 times pmw).

We compared figures indicating the relative incidence of the three diseases in the UK and in the world with the frequencies with which they are mentioned in the OEC. We also compared the annual incidence of these health conditions. Table 2 shows the number of new cases and annual deaths for each disease (note that these figures vary slightly according to different sources).[table 2] The number of new cases of heart failure and dementia in the UK are not dissimilar, at 200,000 and 209,600 respectively, as are the number of annual UK deaths caused by each disease (64,000 and 66,424, respectively; heart failure is included with deaths from coronary heart disease in this figure and dementia is included with deaths from Alzheimer’ s Disease).

**Table 2.** Incidence of heart failure, cancer and dementia in the UK and worldwide

|  | New cases per year |  | Deaths per year |  |
| --- | --- | --- | --- | --- |
|  | UK | Worldwide | UK | Worldwide |
| **Heart failure** | 200,000<sup>1</sup> | 17,900,000<sup>1</sup> | 64,000*<sup>1</sup> | 9,100,000*<sup>1</sup> |
| **Cancer** | 375,400<sup>1</sup> | 17,000,000<sup>1</sup> | 166,533<sup>1</sup> | 9,600,000<sup>1</sup> |
| **Dementia** | 209,600<sup>1</sup> | 9,900,000<sup>1</sup> | 66,424**<sup>1</sup> | 1,500,000***<sup>1</sup> |
| *Coronary heart disease (rather than heart failure specifically)<br>**Includes deaths from dementia and Alzheimer's Disease<br>***Estimated |  |  |  |  |

Table 3 shows the raw and relative frequencies of ‘heart failure’, ‘cancer’ and ‘dementia’ in different genres of contemporary English, according to the OEC text-type classifications (in descending order of raw frequency of ‘heart failure’).

**Table 3.** Comparison of relative frequency of ‘heart failure’, ‘cancer’ and ‘dementia’ in different genres of contemporary English

| Genre (text-type) | ‘heart failure’ |  | ‘cancer’ |  | ‘dementia’ |  |
| --- | --- | --- | --- | --- | --- | --- |
|  | *n* | pmw | *n* | pmw | *n* | pmw |
| Medicine | 7,646 | 102.08 | 62,075 | 828.74 | 3,891 | 51.95 |
| News | 948 | 1.39 | 65,358 | 95.72 | 2,243 | 3.29 |
| Unclassified | 433 | 1.25 | 16,563 | 47.69 | 781 | 2.25 |
| Fiction | 345 | 4.56 | 1,338 | 17.67 | 62 | 0.82 |
| Life and leisure | 194 | 1.44 | 13,938 | 103.53 | 364 | 2.70 |
| Science | 193 | 1.70 | 10,413 | 91.80 | 283 | 2.49 |
| Society | 124 | 1.04 | 5,473 | 45.99 | 160 | 1.34 |
| Weblog | 115 | 0.53 | 7,057 | 32.74 | 318 | 1.48 |
| Arts | 96 | 0.59 | 3,262 | 20.11 | 297 | 1.83 |
| Sport | 83 | 0.79 | 2,474 | 23.62 | 35 | 0.33 |
| Business | 67 | 0.66 | 3,346 | 33.01 | 59 | 0.58 |
| Religion | 19 | 0.43 | 1,843 | 42.10 | 200 | 4.57 |
| Humanities | 21 | 0.43 | 796 | 16.16 | 116 | 2.35 |
| Law | 16 | 0.27 | 524 | 8.72 | 61 | 1.02 |
| Military | 13 | 0.51 | 528 | 20.90 | 1 | 0.04 |
| Computing | 13 | 0.16 | 1,179 | 14.67 | 31 | 0.39 |
| Agriculture | 11 | 0.95 | 677 | 58.60 | 7 | 0.61 |
| Environment | 7 | 0.78 | 1,741 | 194.78 | 18 | 2.01 |
| Paranormal | 4 | 0.68 | 596 | 101.44 | 8 | 1.36 |
| Transport | 2 | 0.19 | 59 | 5.61 | 0 | 0 |
| Games | 0 | 0 | 11 | 2.93 | 10 | 2.66 |
| pmw = per million words |  |  |  |  |  |  |

Unsurprisingly the highest frequencies of all three terms were in the Medical genre, where they were used in biomedical senses in the discussion of medical research. ‘Cancer’ was mentioned about eight times more often than ‘heart failure’ in medical articles, and ‘heart failure’ was mentioned nearly twice as often as ‘dementia’.

Outside of the Medical genre, the words most typically occurring with ‘heart failure (the ‘collocates’) were other medical technical terms in biomedical contexts, revealing nothing of the person’ s experience of heart failure. For example, in the Life and leisure genre the collocates were ‘congestive’, ‘CHF’, ‘hypertension’ and ‘kidney’, and in the News genre ‘congestive’, ‘cardiomyopathy’ and ‘haemorrhage’. Apart from technical terms specifying some aspect of the illness, ‘heart failure’ was also typically associated with the word ‘died’ through the formulaic reporting of heart failure as the cause of death of a well-known person or public figure, as in ‘X (has) died from/of heart failure’. Some examples are shown in the extracts from the corpus data in Table 4.

**Table 4.**
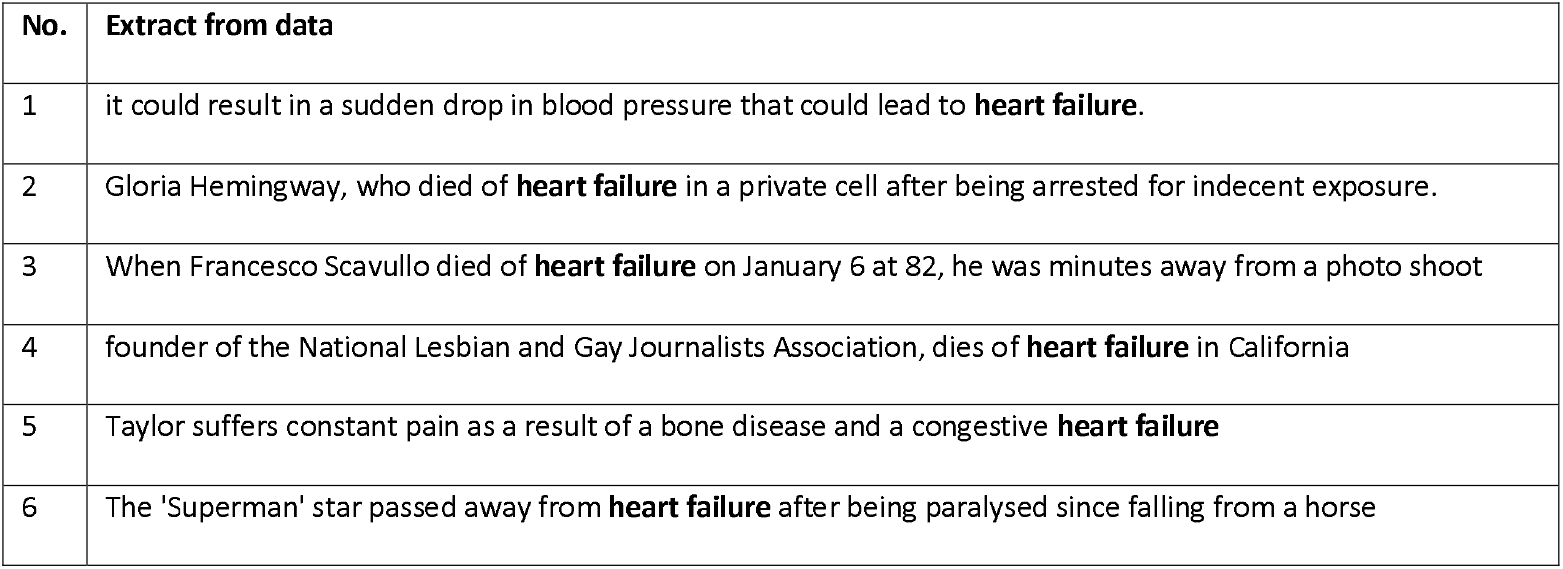
Examples of ‘heart failure’ in OEC Life and leisure genre used to discuss cause of death

| No. | Extract from data |
| --- | --- |
| 1 | it could result in a sudden drop in blood pressure that could lead to **heart failure**. |
| 2 | Gloria Hemingway, who died of **heart failure** in a private cell after being arrested for indecent exposure. |
| 3 | When Francesco Scavullo died of **heart failure** on January 6 at 82, he was minutes away from a photo shoot |
| 4 | founder of the National Lesbian and Gay Journalists Association, dies of **heart failure** in California |
| 5 | Taylor suffers constant pain as a result of a bone disease and a congestive **heart failure** |
| 6 | The 'Superman' star passed away from **heart failure** after being paralysed since falling from a horse |

While other details of the person’ s life emerged in the wider context, the illness of HF itself was not usually discussed except in the context of the death having occurred. In the Life and leisure genre ‘cancer’ was often used in biomedical contexts, but also in contexts more oriented towards people’ s personal and emotional experience of the illness. The collocates of ‘cancer’ in the Life and leisure genre were mainly technical medical terms, as for ‘heart failure’, e.g. ‘incidence’ (n=89), ‘disease’ (n=498), ‘diabetes’ (n=104) and ‘liver’ (n=97). However, in contrast to ‘heart failure’, there were also two person-oriented collocates for ‘cancer’ : ‘survivors’ (n=79) and ‘battling’ (n=50), examples of which are shown in Table 5.

**Table 5.**
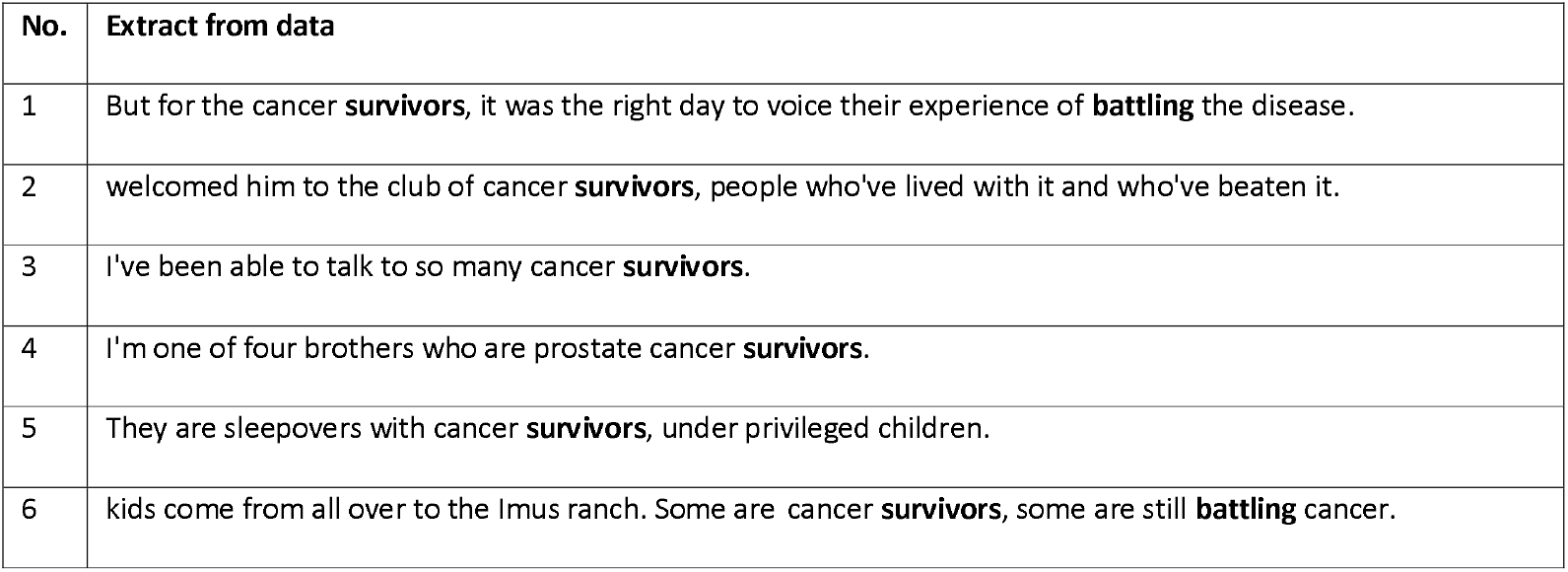
Examples of ‘survivors’ and ‘battling’ associated with ‘cancer’ in contemporary English life and leisure genre

The ‘survivor’ narratives framed people with cancer as being relatively active and empowered. Through the use of violence metaphors ‘battling’ (lines 1 and 6) and ‘beaten’ (line 2), people with cancer were portrayed as actively aiming to live as long as possible, or recover. In line 1, people with cancer were framed as being able ‘to voice their experience’, another potentially empowering activity.

In the Life and leisure genre, there were some cases in which ‘dementia’ was discussed technically in biomedical contexts, shown in lines 1, 3, 4 and 8 in the examples in Table 6.

**Table 6.**
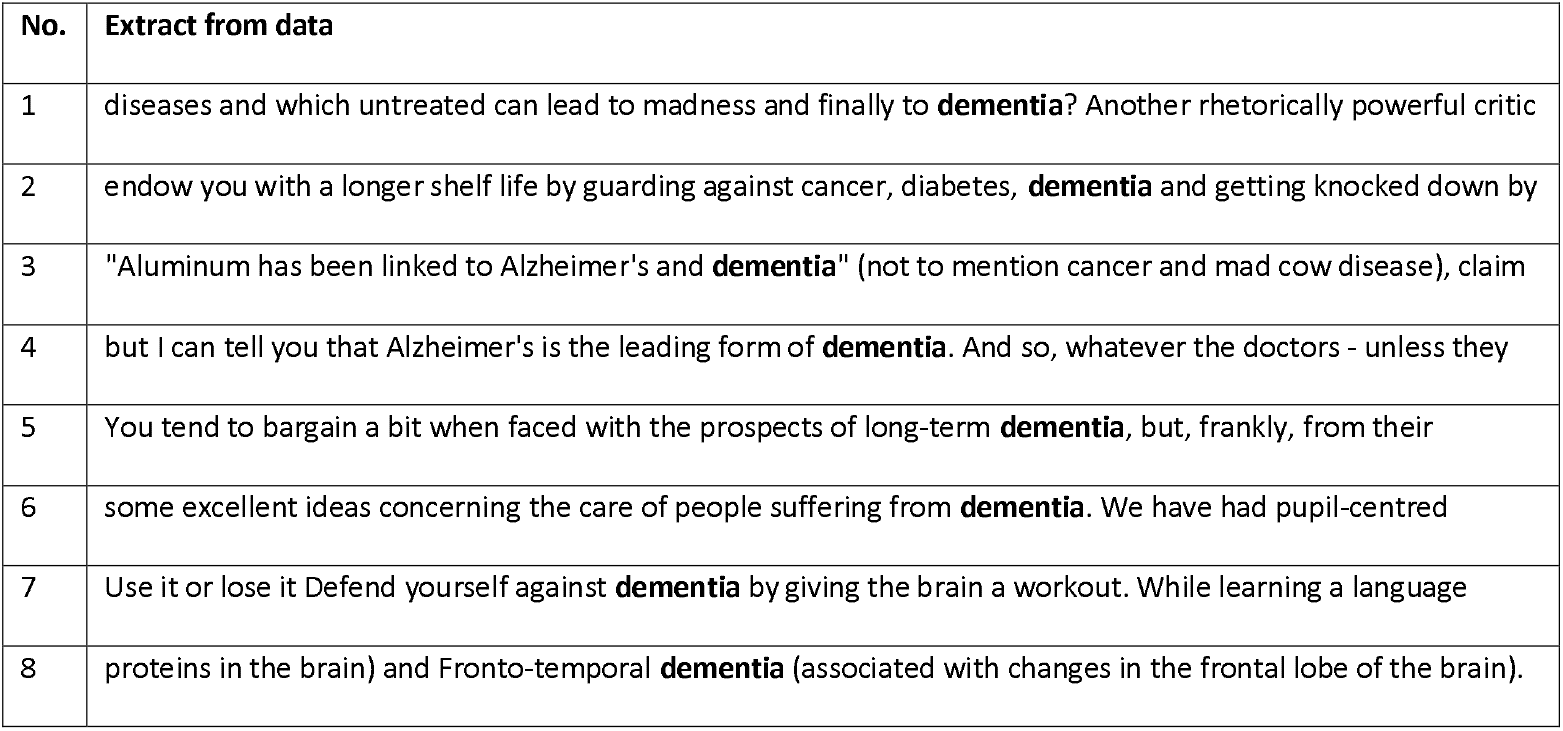
Examples of ‘dementia’ in contemporary English life and leisure genre

| No. | Extract from data |
| --- | --- |
| 1 | diseases and which untreated can lead to madness and finally to **dementia**? Another rhetorically powerful critic |
| 2 | endow you with a longer shelf life by guarding against cancer, diabetes, **dementia** and getting knocked down by |
| 3 | "Aluminum has been linked to Alzheimer's and **dementia**" (not to mention cancer and mad cow disease), claim |
| 4 | but I can tell you that Alzheimer's is the leading form of **dementia**. And so, whatever the doctors - unless they |
| 5 | You tend to bargain a bit when faced with the prospects of long-term **dementia**, but, frankly, from their |
| 6 | some excellent ideas concerning the care of people suffering from **dementia**. We have had pupil-centred |
| 7 | Use it or lose it Defend yourself against **dementia** by giving the brain a workout. While learning a language |
| 8 | proteins in the brain) and Fronto-temporal **dementia** (associated with changes in the frontal lobe of the brain). |

The collocates of ‘dementia’ in the Life and leisure genre were medical/technical terms (‘senile’, ‘Vascular’, ‘Alzheimer’ and ‘colon’). There were examples of ‘dementia’ being used to frame people experiencing it in relatively empowering ways, as for ‘cancer’. For instance, in line 5, Table 6, the writer mentioned bargaining, a process which framed the person with dementia as having some agency and power, although with or against whom was not clear from the wider context (perhaps medical professionals). In line 6 the writer used a violence metaphor to advise the reader to: “Defend yourself against dementia …”. In both scenarios a sense of opposition was created around the use of ‘dementia’, with the first writer indicating there is some sort of deal to be done and the second that the illness is an opposing force. There were no similar cases of such narratives for ‘heart failure’.

### Comparative frequency of use of the terms ‘heart failure’, ‘cancer’ and ‘dementia’ in parliamentary debates from 1945 to early 2021

Figure 1 shows the frequency of use ‘heart failure’, ‘cancer’ and ‘dementia’ in Hansard reports of UK House of Commons and House of Lords debates from 1 January 1945 (including the period leading up to the National Health Services Act of 1946 and the subsequent opening of the NHS in 1948) to 25 February 2021 (the latest date for which data was available).

**Figure 1.**
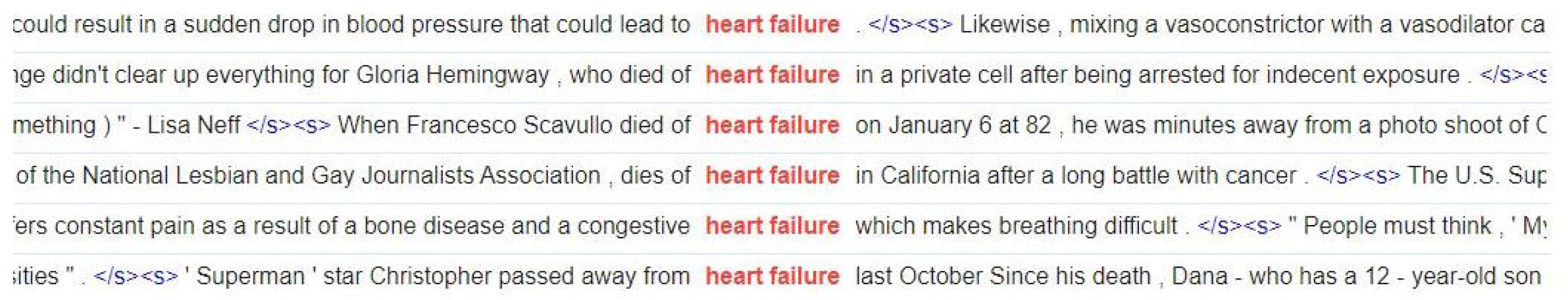
Examples of ‘heart failure’ in OEC Life and leisure genre used to discuss cause of death

The peak frequency of use of ‘heart failure’ in parliamentary debates was just under 1.0 pmw, in 2007.

Figure 1 shows that ‘heart failure’ was used with much lower frequency than ‘cancer’ across the whole time frame, and much lower frequency compared to ‘dementia’ from about 1990 onwards, so much so that the red line on the graph plotting instances of ‘heart failure’ is relatively invisible. Even when compared to a different, non-medical issue of arguably lower importance (in terms of potential threat to human life expectancy and quality of life), potholes in UK roads and pavements, ‘heart failure’ is discussed much less, as shown in Figure 2.

**Figure 2.**
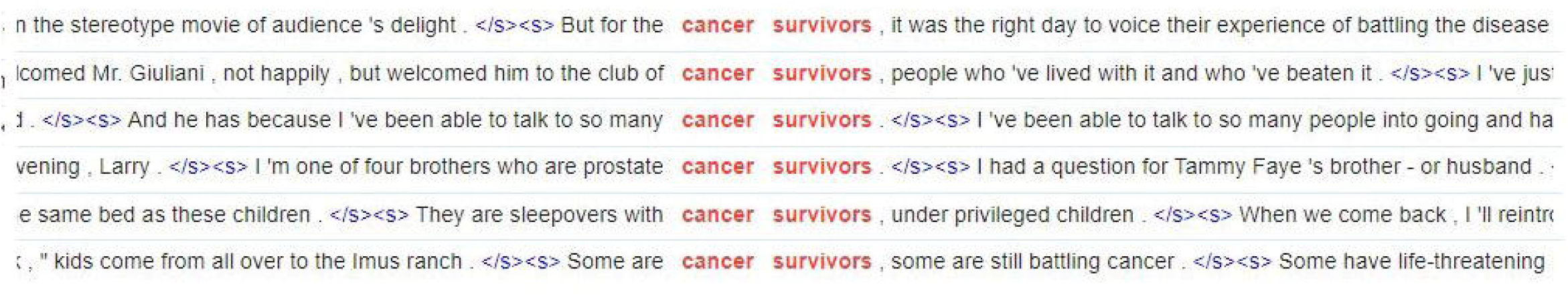
Examples of ‘survivors’ and ‘battling’ associated with ‘cancer’ in contemporary English life and leisure genre

**Figure 3.**
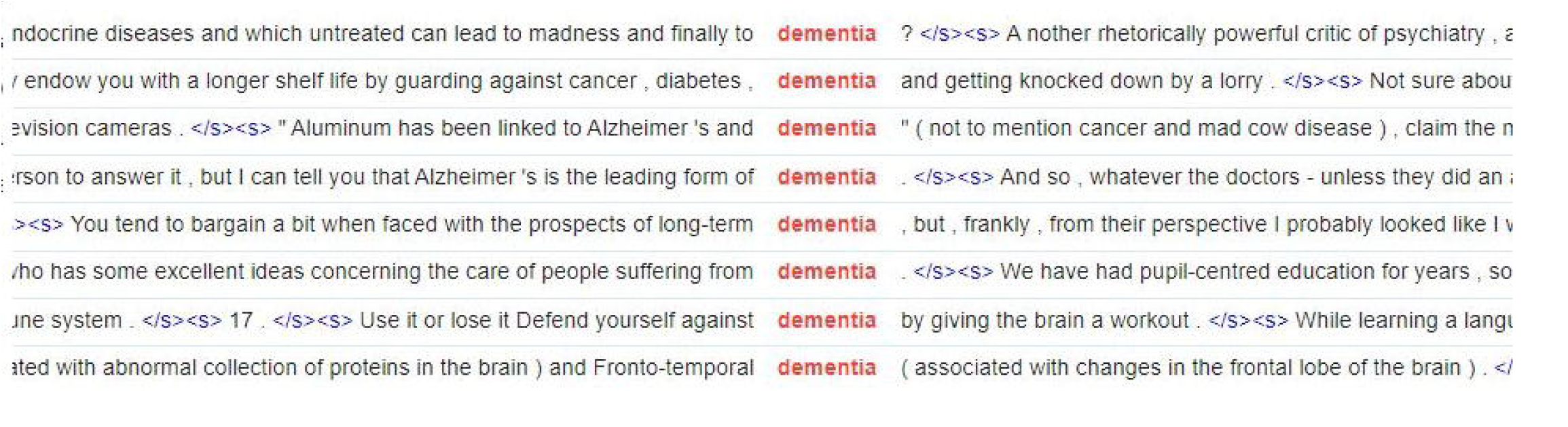
Examples of ‘dementia’ in contemporary English life and leisure genre

**Figure 4.**
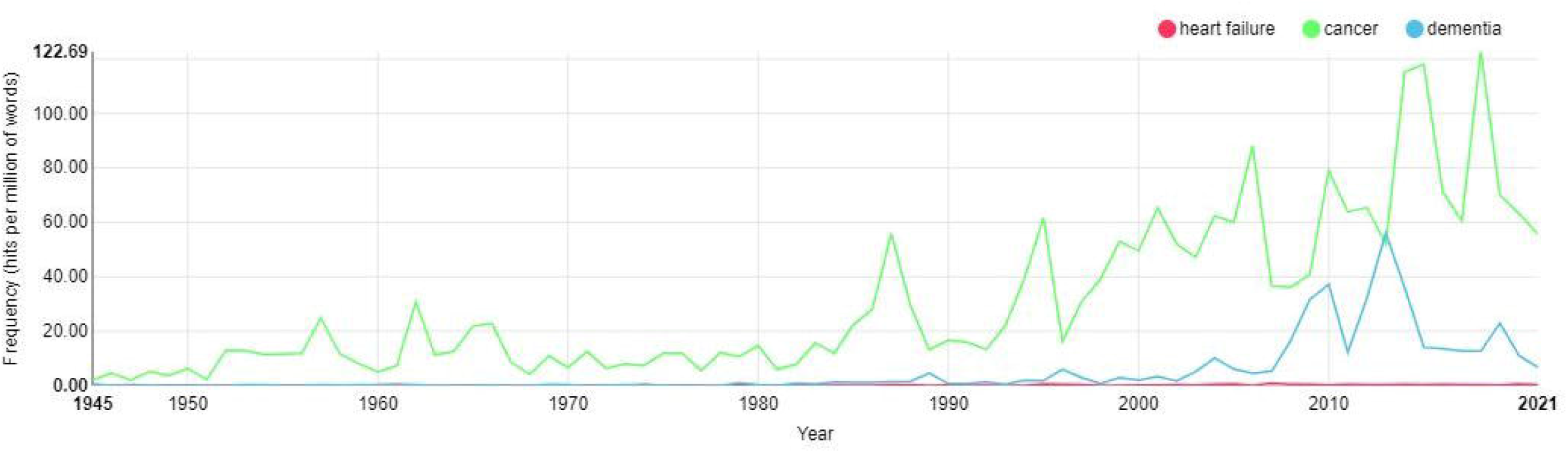
Distribution of ‘heart failure’, ‘cancer’ and’ dementia’ in UK parliamentary debates 01.01.1945 – 25.02.2021

**Figure 5.**
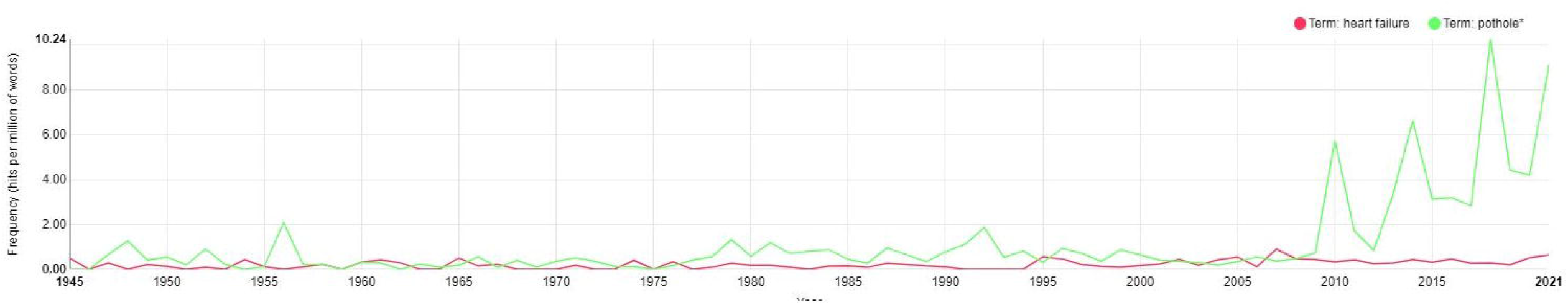
Distribution of ‘heart failure’ compared to ‘pothole/s’ in UK parliamentary debates 01.01.1945 – 25.02.2021

As shown in Figure 2, the frequency of use of ‘heart failure’ was, for most of the period 1945 to 2021, lower than the frequency of talk about ‘pothole/s’, particularly over the last ten years when ‘pothole/s’ peaked in terms of frequency at:

- 10.24 times pmw in 2018 (about 37 times more often than ‘heart failure’ at 0.28 times pmw);
- 6.61 times pmw in 2014 (about 16 times more often than ‘heart failure’ at 0.42 times pmw); and
- 5.74 times pmw in 2010 (about 18 times more often than ‘heart failure’ at 0.32 times pmw). Occasionally prior to 2010 talk about ‘heart failure’ rose slightly above talk about ‘pothole/s’, most recently in 2007 when ‘heart failure’ peaked in use at 0.90 times pmw, about three times as often as ‘pothole/s’ (0.36 times pmw).

## Discussion

The similar number of mentions of ‘heart failure’ and ‘dementia’ therefore roughly reflects a similarity in incidence of these diseases in term of numbers of new cases and annual deaths in the UK. The incidence of cancer is higher than that of heart failure and dementia, with about 1.8 times as many new cases of cancer being diagnosed every year compared to the other two diseases, and more than twice as many annual deaths are caused by cancer than by coronary heart disease (including heart failure) or dementia (including Alzheimer’ s Disease). The relative frequency of use of ‘cancer’ in the OEC data is therefore very much higher than the relative incidence of cancer compared to the other two diseases in the UK.

As shown in Table 2 similar numbers of cases of cancer and heart failure are diagnosed worldwide every year (17-18 million), but only about half as many cases of dementia (just under 10 million). The number of worldwide deaths from cancer and coronary heart disease (including heart failure) is also not dissimilar at 9-10 million, again much higher than the 1.5 million deaths from dementia. The relative frequency of use of ‘cancer’ in the OEC data is therefore again very much higher than the relative incidence of cancer compared to the other two diseases worldwide.

The above comparisons indicate, first of all, that cancer is talked about much more frequently relative to either heart failure or dementia and, secondly, that cancer is talked about with disproportionately high frequency relative to the incidence of the three diseases. O’ Hanlon’ s 2019’ s corpus-assisted comparison of Twitter posts concerning breast cancer and heart disease similarly showed that there was much less talk about heart disease than breast cancer, although heart disease was responsible for many more annual deaths (in the US) than breast cancer^15^.

Violence metaphors, especially ‘battle’, ‘struggle’ and ‘fight’, regularly contribute to the construction of vivid scenarios in which people with cancer are relatively empowered. The framings of people with cancer as being ‘survivors’ who are ‘battling’ showed more of a person-centred focus, with vivid descriptors orienting the reader to the person’ s feelings as well as to their behaviours as a cancer sufferer. These contrasted with the formulaic uses of ‘heart failure’ as a cause of death, in which the person who suffers it was framed as a passive recipient. Even in more socially-oriented types of text, talk about HF is mainly of a biomedical nature, used in relatively technical and formulaic ways, especially in reporting cause of death. However, cancer is more typically mentioned in the context of incidence, diagnosis, cure or awareness - in many ways an opposite framing.

In contrast to HF, cancer discussions regularly incorporate figurative language through which people with cancer are framed as ‘survivors’ actively ‘battling’ their illness. Empowering framings of people engaged actively in opposition to heart failure do exist, but these are very much less typical than in discussions of cancer. There is little evidence of person-centred discussion about the experience, feelings and/or emotions of people with HF or their quality of life. Our findings show some similarity to those of O’ Hanlon 2019, who found that talk about heart disease, was less focused on personal experience than talk about breast cancer^15^. The importance of appropriate language has been emphasised recently, particularly in North America where there has been a trend to veer away from using the term ‘failure’ (due to the associated negative connotations) and instead to refer to ‘heart function’ ^23^. However this strategy can also risk a suggestion of appearing to minimise the severity of the condition as there is also evidence that some people with the condition can underestimate how sick they truly are^24^.

If we take frequency of mentions as an indicator of importance, topic of HF has been much less important in UK parliamentary debates in recent years than even potholes in roads and pavements. Whether this reflects the priorities of the parliamentarians, their constituents, or both, we cannot say from the information available. It is possible that more constituents contacted their elected representatives to complain about pot-holes than about provisions for the treatment and support of HF. We should note that, in addition to general frustration and inconvenience, pot-holes do pose some threat to health and quality of life (the AA reported in 2018 that 22 cyclists were killed and 368 seriously injured from accidents caused by potholes over a 12-month period^25^). We might speculate that the amount of (negative) UK media coverage regarding pot-holes could be greater than that for HF, which may result in greater amounts of concern expressed in parliamentary debates.

It would be possible to investigate words most typically associated with ‘heart failure’ and ‘cancer’ on a statistical basis using the Wordsketch tool in SketchEngine, which identifies collocates according to grammatical function. This can be useful because words with different grammatical functions have particular roles. For instance, nouns are used for naming (e.g. cancer ‘survivor/s’), and they function as subjects or objects (i.e. as social actors/agents who carry out actions or who are the recipients of actions). Verbs describe states, actions and processes, including what is being done or experienced (e.g. ‘heart failure’ typically occurs with verbs ‘die’ and ‘suffer’). Investigating the grammatical characteristics of words typically occurring in the context of ‘heart failure’ and ‘cancer’ could potentially reveal more details about the situations and framings in which the illnesses tend to occur.

It would also be possible and potentially useful to conduct a larger study using a wider range of cardiovascular terms (e.g. ‘heart attack’, ‘cardiovascular disease’, ‘CHF’) and to compare their frequency and manner of use with those for other health conditions apart from cancer, for example dementia/Alzheimer’ s Disease.

### Limitations

It was outside the scope of this study to compare the frequency and manner of use of ‘heart failure’ and ‘cancer’ in different geographical varieties of contemporary English in detail (aside from noting overall trends in relative frequencies above). It was also not possible to discuss the use of ‘heart failure’, ‘cancer’ and ‘dementia’ in every genre of contemporary English, so we have reported on those showing the greatest contrasts.

A debate on ‘Patients with Heart Failure’ took place in the UK House of Commons on 11 March 2021, slightly later than the most recent debates currently accessible through the HC corpus interface, so it is not included in our data. The transcript is available at https://hansard.parliament.uk/Commons/2021-03-11/debates/14BCE210-9636-4060-8D0B-31D9425BD334/PatientsWithHeartFailure?highlight=heart%20failure#contribution-98D83586-DFF2-4926-A7A7-2B0FA62A831E.

## Conclusion

Our study has elucidated that heart failure is relatively under-discussed in comparison to other conditions such as cancer and dementia, both in societal discourse as well as in UK parliamentary debates. Despite comparable morbidity and mortality, discussions regarding people with HF are less person-centred and empowering in comparison to the language used to describe people with cancer. In UK parliamentary debates, HF is also talked about less frequently than non-medical topics such as pot-holes, which, although non-trivial, are arguably less important and urgent. It is crucial that all stakeholders involved in HF redouble their efforts to spread awareness regarding the seriousness of the condition and the pressing need to significantly improve investment in prevention, early diagnosis and better management.

## Data Availability

All data produced in the present study are available upon reasonable request to the authors

## Ethics

No data was collected and analysed involving human or animal participants for this study.

## upporting information

None.

